# Improving the reproduction number calculation by treating for daily variations of SARS-CoV-2 cases

**DOI:** 10.1101/2021.08.15.21262071

**Authors:** Harry Drewes, Gotthold Fläschner, Peter Möller

## Abstract

The Covid-19 pandemic impacted the human life all over the globe, starting in the year of its emergence, 2019, and in the following years. A epidemiological key indicator that gained particular recognition in politics and decision making is the time-dependent reproduction number *R*_*t*_, which is commonly calculated by institutions responsible for disease control following a method presented by Cori et. al. Here, we propose an improved as well as an alternative method, which make the calculation more stable against oscillations arising from daily variations in testing. Both methods can be used without great statistical knowledge or effort. The methods provides a smoother result without increasing the time-lag, and provides an advantage particular in the timeframe of weeks, which might serve as a better ground for forecasts and the raising of alarms.

## INTRODUCTION

The *R*_*t*_-value describes the average number of people an individual is expected to infect and is, therefore, acting as a measure of transmissibility and can provide feedback on the effectiveness and need of interventions. Particularly during the Covid-19 pandemic, this measure gained recognition, albeit experts warned to not exclusively focus on it, as it does not account for all the complex dynamics during a pandemic^1^. Nonetheless, the *R*_*t*_-value provides an important and easy to grasp concept, which aids political decision making. As such an *R*_*t*_ > 1 indicates an acceleration of the pandemic and, conversely, an *R*_*t*_ < 1, leads to a slowing-down. Therefore, political decision makers are anxious to keep *R*_*t*_ below 1, and tighten regulations otherwise. Different methods to calculate the *R*_*t*_-value have been developed to varying degrees of complexity, depending on the requirements (e.g., “ease-of-use” and statistical information). Cori et. al. presented a method of particular use for non-experts of pandemic model building, as it is easy to implement and robust in its application. The calculation is based on the incidence time series and the serial interval, which is the time offset between the symptoms of a primary case and a secondary case. This approach is prominently applied by the Robert-Koch-Institute^2^ (RKI), the German federal government agency responsible for disease control, but also by the Swiss Government^3^. In case of the RKI, the calculation is based on the now-casting^4^ numbers which estimate the progression of the number of Covid-19 infections and provides a supression of oscillations caused by the reporting delays. However, in many databases and for many countries this kind of now-casting data is not available. In all cases, these data is subject to noise which is why two different *R*_*t*_-values are distinguished by the RKI, depending on different smoothing intervals of *τ* days. The smoothed reproduction number *R*_*t,τ*_ is calculated as^2^:

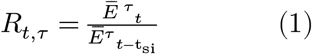

where

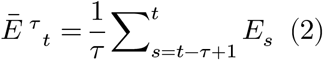

is the over *τ* days averaged number of new infections *E*_s_ on day *s* and *t*_si_ = 4 in case of Cov-Sars2 virus. Typically, an averaging interval of *τ* = 7 is being used. A great advantage of this method - to which we will refer as ‘Cori’s method’ - is, that it can easily be implemented even with spread sheets. We will show (1) that our different approach provides better results in terms of robustness against periodic oscillations and (2) that this approach can be transferred to the standard method by changing the arithmetic mean to a geometric mean, as shown at the end of the results section.

## MATERIALS AND METHODS

### Creation of an R-value test function

We created an *R*_*t*_-value test-function (see Figure 1 and Supplementary Information) to illustrate the differences of the *R*_*t*_-value calculation methods, and generated the curve of hypothetical new-infections 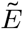 per day, based on the formula

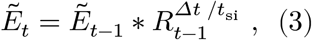

**Figure 1.**
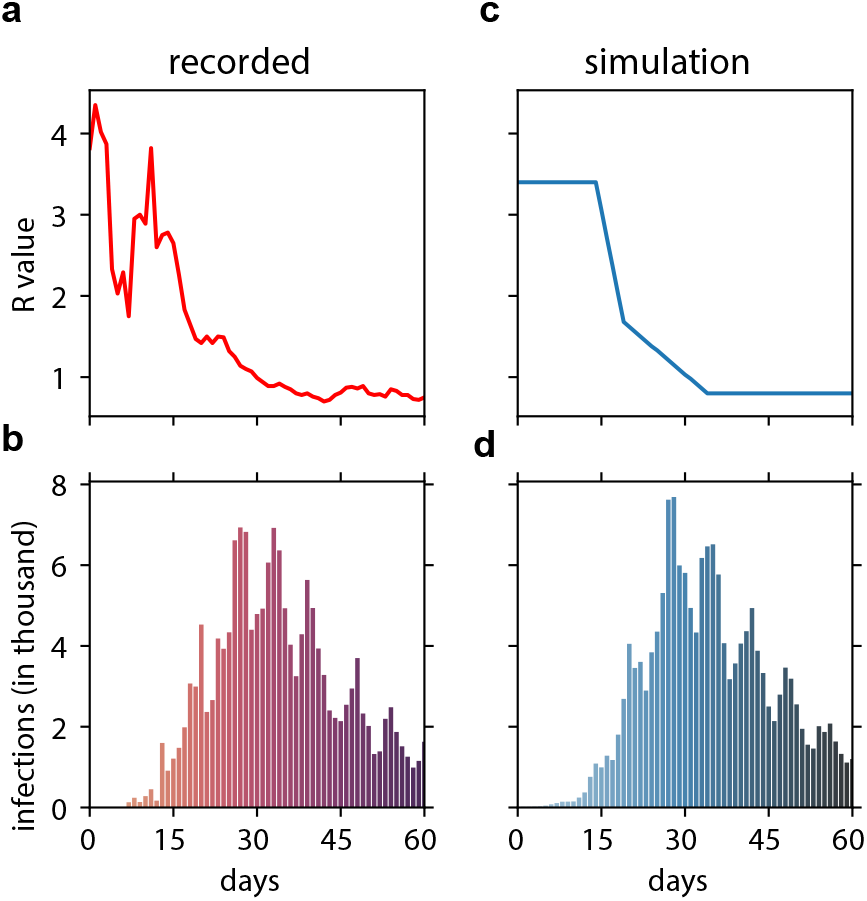
Generation of a test function to compare different extraction models of *R*_*t*_. a *R*_*t*_ as extracted per Cori’s method from based on the data shown in b. b daily infections in Germany in the timeframe from 02.03.2020 to 29.04.2020 as retrieved from the humanitarian data exchange. c *R*_*t*_-value test function used to generate the infection events shown in d. d Simulated infection events based on c. The noise introduced is given as in the materials section.

where Δt = 1 day and the serial interval is t_si_ = 4 days. Eq (3) is yielded from rearranging the definition of 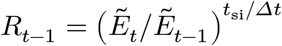. Furthermore, we introduce a multiplicative random noise term (1 + *z*_*t*_) and a multiplicative term that introduces cyclical variations (1 + *d*_*t*_), to mimic variations in daily reporting and medical diagnostics^5^, and write for the number of reported infections 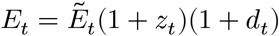. Hereby, z_t_ is a value drawn from a random distribution with mean 0 and a standard deviation *σ*= 0.1, and *d*_*t*_ = *A* cos (2*πt*/*n* − *φ*), where *A* is the amplitude chosen to be 0.25, 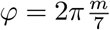 representing a phase offset such that the maximum value of the cosine is found on the *m*th day of the week, and *n* determining the periodicity of the cyclic oscillations, which we found to be 7, as explained by the length of the week. All parameters have been chosen such that *E*_*t*_ mimics the data from the humanitarion data exchange6 for Germany during the time from 02.03.2020 to 29.04.2020, see Fig 1.

### Extraction of *R*_*t*_-value

To extract the *R*_*t*_ we rewrite Eq. 3 by taking the logarithm, such that

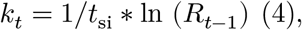

where

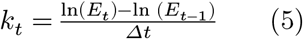

is the slope of a linear equation for a constant *R*_*t*−1._ To find the τ-days averaged 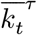, we can minimize the sum of squared deviations (SSD) of τ data points with respect to a straight line. We used *τ* = 7, as it is used by the RKI, resulting in 11 datapoints per one calculation due to the 4 days serial interval. From these 11 datapoints of infectious events, we made two subsets, data ponts 1 to 10 and data ponts 2 to 11, and determined the respective slopes by minimizing the SSD according to the standard expressions of linear regression. See Fig. 2a for an illustration. Subsequently, we averaged the two slopes 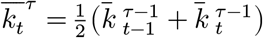 This procedure serves as an addtional mean of noise reduction. From the found 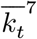, we calculated 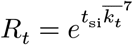

**Figure 2.**
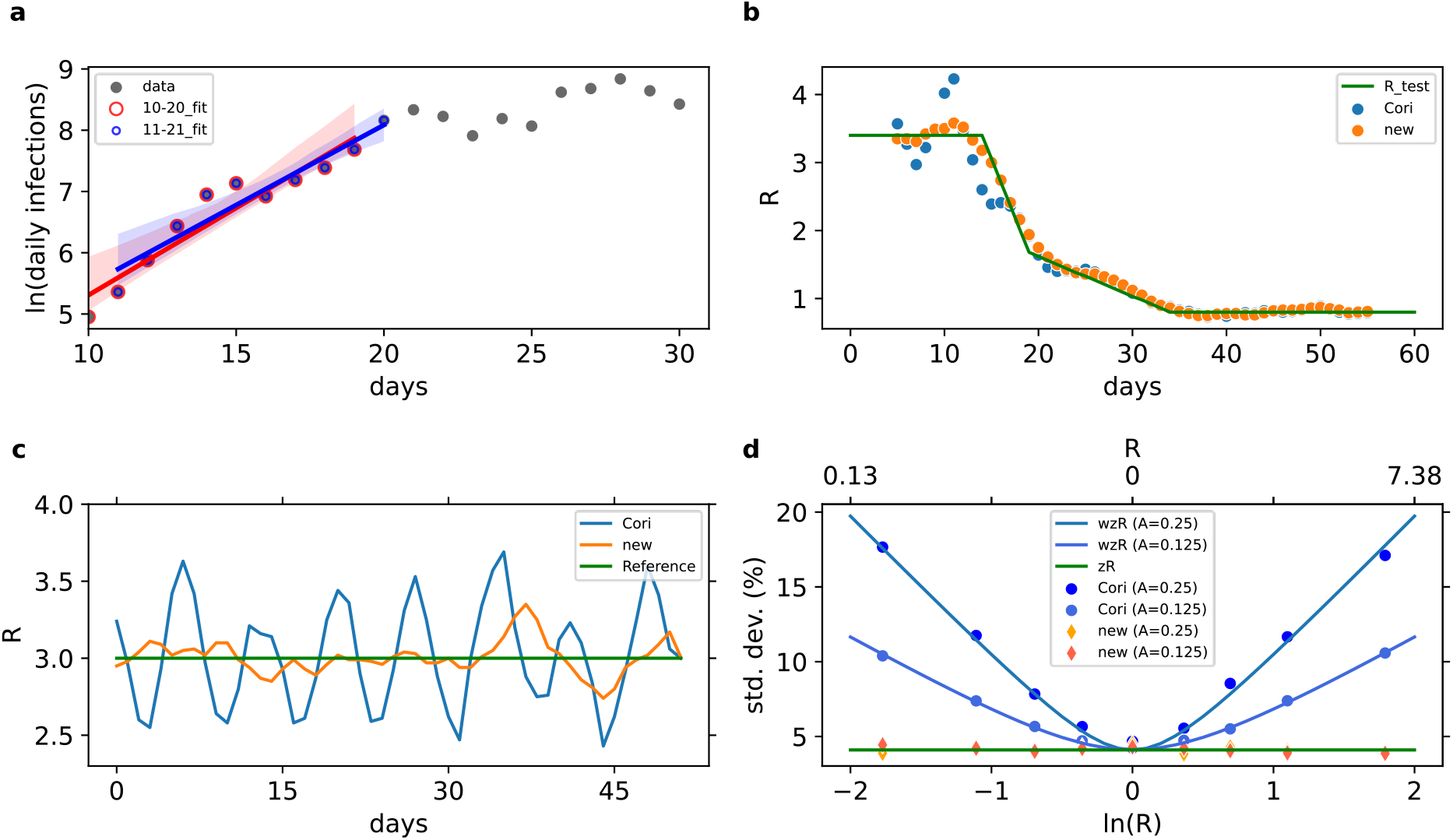
The daily variations in reported infections has a lower impact on the *R*_*t*_ value calculations based method than this of Cori. a The data shown in Fig 1c in the time frame from 10 to 31 days is logarithmically plotted. Linear fits for of 10 days, displaced by one day, are used to calculate the average slope in the center day (in this case day 15). b Extracted *R*_*t*_-values based on a (orange dots) compare better agains Cori’s method (blue dots) in the region of high R_t_-values. c Results of a simulation where the R_t_-value is kept constant at 3 (reference). It is clearly visible, that the daily oscillations can be suppressed by our method (shown in orange). d Dependency of the error of the *R*_*t*_-values for both extraction methods described as standard deviation from the true value. The base line of the error (*zR*) is given by the random noise, whereas the error resulting from random noise and the daily variations (*wzR*) is big for small and big *R*_*t*_-values in Cori’s method (shown in blue). Our method by contrast is not affected (orange, red) by the amplitude *A* of the daily variations.

## RESULTS

The starting point for our simulation comparing the models is seen in Fig. 1a. The infection starts out with a big *R*_*t*_ -value, that is gradually declining as measures to prevent the spread of the disease are taken and is finally dropping to values below 1. The *R*_*t*_ -values are generated based from the original data of the humanitarian data exchange seen in Fig. 1b using Cori’s method. The simplified R_t_ curve which we used as a test-function for comparing ours and Cori’s method is shown in Fig. 1c. It generates infection events (see Materials and Methods) as shown in Fig. 1d, approximating the original data very well. Here, and also for the later deliberations, it is irrelevant that the test-function is not smooth. It is, however, handy to keep the test function that way, as it reduces the number of parameters to generate it.

We took the infection events as shown in Fig. 1d and used them to extract the corresponding R_t_-values using Cori’s method (see Introduction) and our approach (see Materials and Methods). The fits upon which the latter one relies, are in shown in Fig. 2a. As can be seen in Fig. 2b Cori’s method shows strong deviations from the test-function particularly during the times of high *R*_*t*_*-*values. The daily variation is still strong when the averaging over 7 days, during with the infection events are changing a lot. As can be seen in Fig. 2b, the from Cori’s method resulting deviations from the ‘real’ *R*_*t*_-value are considerably big, when either *R*_*t*_ *> 1* or *R*_*t*_ *< 1*, whereas the deviations are small when *R*_*t*_ ≈ 1. However, our model follows the test-function much closer. To further investigate the dependence of the maximum error on our simulation parameters, in particular the noise terms, we varied the standard-deviation σ of the added random noise, as well as the amplitude *A* of the cyclic noise term (see Materials and Methods) and performed 1000 simulations for different *R*_*t*_ values (example shown in Fig. 2c) for given sets of *σ* (0.1) and *A*. (0.25 and 0.125) We calculated the relative error (RE), the results are shown in Fig. 2d. As can be seen, *σ* acts as an additive constant *z*_*R*_, or floor to RE, such that *z*_*R*_ = 0.41*σ*, for both models. For *A* = 0.25, which closely mimicks the behavior seen in the real data, the contribution of the cyclic noise is only found for the Cori’s method and can be described as *w*_*R*_ = 0.38*A* * |ln *R*|. The total error wz_R_ for the Cori’s method is, therefore, 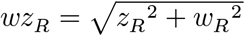, whereas for our method wz_R_=z_R_. This result implies that our method does not suffer from inaccuracies stemming from the daily oscillations.

Next, we explored whether a simple fix could be applied to Cori’s method, starting from understanding the different treatment of the noise contributions in each model. Hereby, we found that due to our effective averaging of the infection data on the level of logarithms, it would be interesting to modify Eq. 2 to be geometric instead of an arithmetic mean. The reasoning relates to the geometric mean expressible as the arithmetic mean of logarithms. We thus modified Eq. 2 and inserted into Eq. 1:

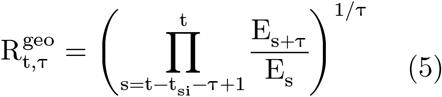

With Eq. 5 we achieved very similar results as with our model, as can be seen in Fig. 3.

**Figure 3.**
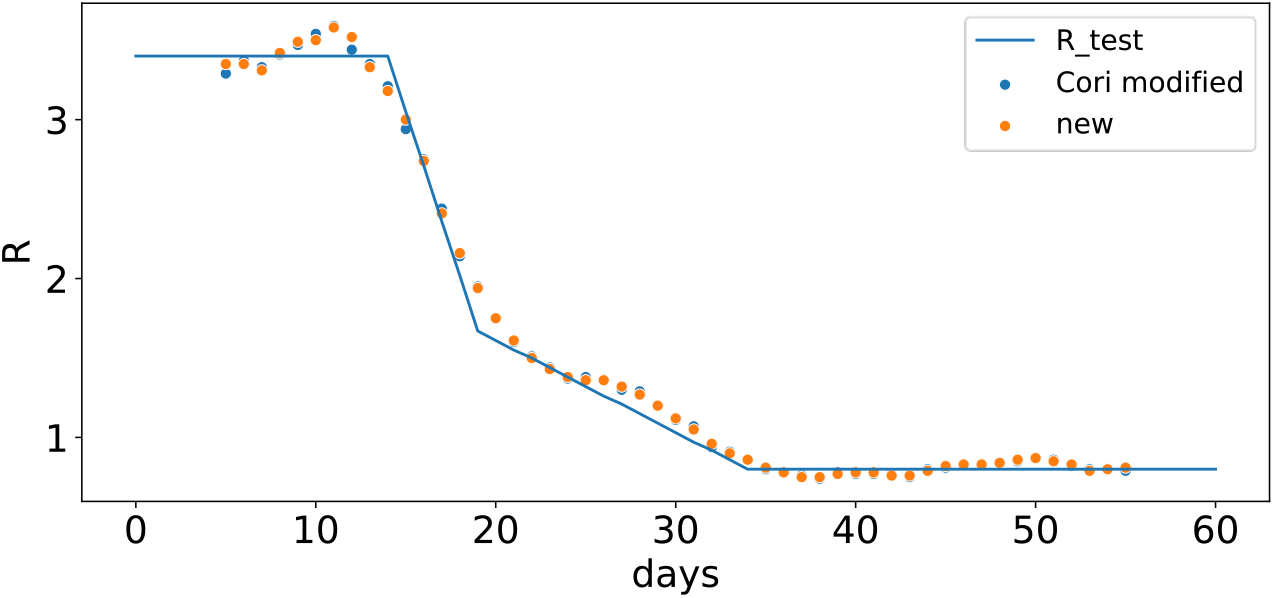
Replacing the arithmetic mean by a geometric mean yields similar results for Cori’s method as by our method based on fitting the slopes of subsets of the infection time series. The daily variations are effectively suppressed.

## DISCUSSION

We have introduced a simple method to determine the R_t_ value based on an approach that is stemming from the control of dynamic systems. In brief, it determines R_t_ as the slope of a linear equation that is a subset of the time series of new infections described in the logarithmic scale. This approach accounts for the exponential nature of the viral spread and is easily implementable via spread sheets. We found that this approach is not susceptible for the daily variations of the nowcasting data as introduced by variations in daily reporting and medical diagnostics as we found it to be the case for Cori’s method for R_t_ > 1 and R_t_ < 1. We further found, that Cori’s method can improved by using geometric averages instead arithmetic averages. As in other applications of finance^7^ and social science^8^, the geometric mean proves to be better to describe growth rates as can easily be illustrated by the compound annual growth rate^9^: an initial growth of 80% and a subsequent growth of 25% for instance, is effectively an average growth by 50%, and not 52.5% as derived by the arithmetic mean. To conclude, we believe that the simple change from arithmetic to geometric mean or alternatively our method might prove a valuable tool to determine the R_t_-values on time frames where cyclical variations are present - as in the days of the week - in particular, when the occurrence of infections is changing rapidly. This in turn, is beneficial to avoid false alarms and to strengthen the trust of the population in the data extracted and therefore in the governmental organizations and scientists, as the data is visibly more free of noise. It can also be of importance for making better forecasts.

## Data Availability

The underlying data sets are publically available.
The generation of the remaining datasets is explained in the text, they are however available upon request.

https://data.humdata.org/dataset/novel-coronavirus-2019-ncov-cases

## ACKNOWLEDGEMENTS

The authors want to thank Claudius Noack for the fruitful discussions. This work was supported by Hamburg University of Applied Sciences.

## REFERENCES

1 Adam, D. A guide to R - the pandemic’s misunderstood metric. Nature 583, 346–348, doi:10.1038/d41586-020-02009-w (2020).

2 Erläuterung der Schätzung der zeitlich variierenden Reproduktionszahl R, Robert Koch Institut Berlin, (2020).

3 Huisman, J. S. et al. Estimation and worldwide monitoring of the effective reproductive number of SARS-CoV-2. medrxiv (2020).

4 Hohle, M. & an der Heiden, M. Bayesian nowcasting during the STEC O104:H4 outbreak in Germany, 2011. Biometrics 70, 993–1002, doi:10.1111/biom.12194 (2014).

5 Bergman, A., Sella, Y., Agre, P. & Casadevall, A. Oscillations in U.S. COVID-19 Incidence and Mortality Data Reflect Diagnostic and Reporting Factors. mSystems 5, doi:10.1128/mSystems.00544-20 (2020).

6 https://data.humdata.org/dataset/novel-coronavirus-2019-ncov-cases

7 Rowley, E. E. The Financial System Today. (Manchester University Press, 1987).

8 index, h. d. Why is the geometric mean used for the HDI rather than the arithmetic mean?, http://hdr.undp.org/en/content/why-geometric-mean-used-hdi-rather-arithmetic-mean (2021).

9 J. P. Anson, M. J. Fabozzi F. & J. Jones F. The Handbook of Traditional and Alternative Investment Vehicles: Investment Characteristics and Strategies. (John Wiley & Sons, 2010).

